# Analysis of Covid-19 and non-Covid-19 viruses including influenza viruses to see the influence of intensive preventive measures among Japanese

**DOI:** 10.1101/2020.05.21.20105106

**Authors:** Yosuke Hirotsu, Makoto Maejima, Yumiko Kakizaki, Yoshihiro Miyashita, Hitoshi Mochizuki, Masao Omata

## Abstract

Severe acute respiratory coronavirus 2 (SARS-CoV-2) spread and cause death in worldwide. The preventative measures and infection control are underway throughout the society and there are signs of convergence in some areas. Other viruses as well as SARS-CoV-2 cause cold-like symptoms and spread in winter. However, it is unclear to what extent SARS-CoV-2, influenza virus and other causative viruses have been prevailed since the preventative measures were implemented. In this study, we conducted multiples PCR and quantitative reverse transcription PCR using nasal swabs from 191 patients with cold-like symptoms in Japan to reveal the causative viruses. As a result, at least one virus were detected in 40 out of 191 (21%) patients. Of these, we frequently identified the human rhinovirus / enterovirus (5.8%, n=11), SARS-CoV-2 (4.2%, n=8) and human metapneumovirus (3.7%, n=7). On the other hand, no influenza virus was detected. These results shows the prevalence of causative viruses after the social preventative measures and implies the difference of infectivity between SARS-CoV-2 and influenza virus.

## INTRODUCTION

The new emergent coronavirus, severe acute respiratory coronavirus 2 (SARS-CoV-2), broke out in Wuhan, China [1]. The virus immediately spreads around the world and causes a huge number of COVID-19 and deaths. SARS-CoV-2 presents in the upper respiratory tract and spreads by droplets from coughing or sneezing. World Health Organization recommends to clean hands with soap, keep social distance, and avoid going to crowded places in order to prevent the spread of viral infections [2]. In addition to these preventative measures, each country recommends travel restrictions and curfews.

Symptoms like the common cold are caused by some viruses including influenza virus, rhinovirus, adenovirus, coronavirus (229E, OC43, NL63 and HKU1) and human metapneumovirus [3]. Of these, influenza virus is an annually epidemic in the winter. Owing to the development of treatments and vaccine, its mortality rate has become to be low.

In December 2019, outbreak of SARS-CoV-2 just coincides with the epidemic of influenza virus. Seasonal influenza activity was lower in 2020 than in previous years in Japan [4]. However, it remains unknown whether SARS-CoV-2 or other virus is more strongly infectious. Here, we performed genetic analysis to determine which virus is spreading in the population in Japan. This epidemiologic study shows the infectability of each virus after taking social preventive measures against infectious diseases.

## METHODS

We conducted the cohort study to 191 patients with cold-like symptoms from 10 March 2020 to 7 May 2020 in our district distant from 100-150 km west of Tokyo, Japan. Nasopharyngeal swabs were collected from all patients and subjected to subsequent genetic analysis. To analyze the virus species related to respiratory diseases, we performed multiplex PCR targeting 17 viruses and three bacteria using FilmArray Respiratory Panel (bioMérieux, Marcy-l’Etoile, France) (Table 1). We performed real-time quantitative reverse transcription PCR (RT-qPCR) to detect SARS-CoV-2 as previously described [5].

**Table 1.** The number of patients infected with SARS-CoV-2 and respiratory infection virus (n=40)

| **Viruses** | **n** |
| --- | --- |
| Human Rhinovirus / Enterovirus | 11 |
| SARS-CoV-2* | 8 |
| Human Metapneumovirus | 7 |
| Coronavirus 229E | 4 |
| Coronavirus OC43 | 3 |
| Adenovirus | 2 |
| Respiratory syncytial virus | 2 |
| Coronavirus NL63 | 1 |
| Adenovirus and Human Rhinovirus/Enterovirus | 1 |
| Adenovirus and Human Metapneumovirus | 1 |
| Coronavirus HKU1 | 0 |
| Influenza A | 0 |
| Influenza A/H1 | 0 |
| Influenza A/H1-2009 | 0 |
| Influenza A/H3 | 0 |
| Influenza B | 0 |
| Parainfluenza 1 | 0 |
| Parainfluenza 2 | 0 |
| Parainfluenza 3 | 0 |
| Parainfluenza 4 | 0 |
| **Bacteria** |  |
| *Bordetella pertussis* | 0 |
| *Chlamydophila pneumoniae* | 0 |
| *Mycoplasma pneumoniae* | 0 |
| **Total** | **40** |

SARS-CoV-2, severe acute respiratory coronavirus 2
\*SARS-CoV-2 were detected by RT-qPCR

The Institutional Review Board at Yamanashi Central Hospital (YCH) approved this study and the use of an opt-out consent method. The requirement for written informed consent was waived. Patients had the opportunity to refuse to participate in the study.

## RESULTS

Of 191 patients, 32 (17%) were infected at least one causative virus. There were human rhinovirus / enterovirus (n=11), human metapneumovirus (n=7), coronavirus 229E (n=4), coronavirus OC43 (n=3) and adenovirus (n=2), respiratory syncytial virus (n=2), coronavirus NL63 (n=1) (Table 1). Two patients were infected with two virus coincidentally, including adenovirus and human rhinovirus / enterovirus (n=1), adenovirus and human metapneumovirus (n=1) (Table 1). However, influenza A, influenza A/H1, influenza A/H3, influenza A/H1-2009 and influenza B viruses were not detected. These results suggested influenza virus was very low prevalence in our district or prevented by intensive measures taken against coronavirus.

In the same cohort, we conducted real-time RT-qPCR to detect SARS-CoV-2. Among of 191 patients, 8 were infected with SARS-CoV-2 and all of these were observed in patients who were not infected other causative virus by FilmArray Respiratory Panel (Figure 1). Thus, the prevalence rate of SARS-CoV-2 infection was 4%, but that of influenza virus was 0% in 191 patients (Figure 1). This comparative analysis suggested the infectability of SARS-CoV-2 was much higher than that of influenza virus under the same circumstances and same region.

Collectively, the respiratory panel could reveal as many as 17% (32/191) of the causative virus detect 7 types of virus species; whereas no influenza virus was detected. At the very beginning of the epidemic of coronavirus, there was a thought that the infectivity of coronavirus could be no more than that of influenza.

**Figure 1.**
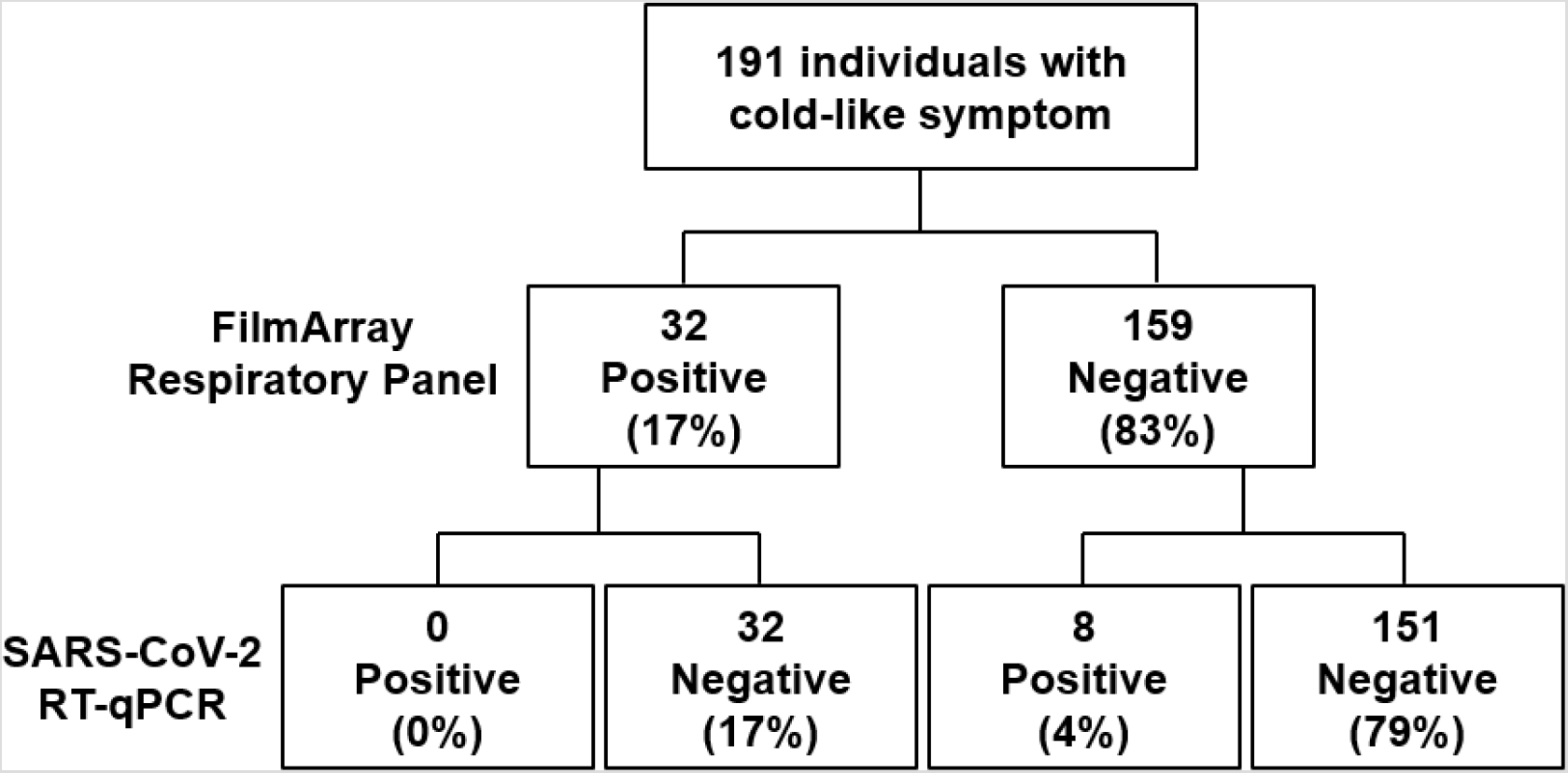
Virus infection status in 191 patients. **(A)** Flow chart shows the number of patients who was infected with causative virus. FilmArray Respiratory Panel detected the 32 positive and 159 negative patients. RT-qPCR analysis detected SARS-CoV-2 in 8 patients.

## DISCUSSION

This study implied the difference of infectivity between SARS-CoV-2 and influenza virus. The infection prevention measures such as handwashing and wearing a mask led to a decrease in influenza infections [6]. Especially, the latter had been a common practice among Japanese in winter time.

Although prospective and updated, the limitation was that this single institution study was conducted in our district. Our national registry indicated marked reduction of influenza virus infection not in beginning, but in the latter time of this winter which coincided with our study. Further studies will be necessary to verify whether the lower infection prevalence of influenza transmission by social preventive measures.

We learned a lesson that by taking the stringent measure we may prevent influenza viruses which have had bigger impact on human life for long.

## Data Availability

The data is available upon request.

## Acknowledgement

We thank all of the medical and ancillary hospital staff and the patients for consenting to participate.

